# Implementation of Community Health Worker Support for Tobacco Cessation: A Mixed-Methods Study

**DOI:** 10.1101/2024.01.26.24301835

**Authors:** Cheryl Y. S. Foo, Kevin Potter, Lindsay Nielsen, Aarushi Rohila, Melissa Culhane Maravic, Kristina Schnitzer, Gladys N. Pachas, Douglas E. Levy, Sally Reyering, Anne N. Thorndike, Corinne Cather, A. Eden Evins

## Abstract

**Objective:** Adults with serious mental illness have high tobacco use disorder rates and underutilization of first-line tobacco cessation pharmacotherapy. In a randomized trial, participants offered community health worker (CHW) support and primary care provider (PCP) education had higher tobacco abstinence rates at two years, partly through increased tobacco cessation pharmacotherapy initiation. This study determined the association between participant-CHW engagement and tobacco abstinence outcomes.

**Methods:** This was a secondary, mixed-methods analysis of 196 participants in the trial’s intervention arm. Effects of CHW visit number and duration, CHW co-led smoking cessation group sessions attended, and CHW-attended PCP visit number on tobacco use disorder pharmacotherapy initiation and tobacco abstinence were modeled using logistic regression. Interviews with 12 CHWs, 16 participants, and 17 PCPs were analyzed thematically.

**Results:** Year-two tobacco abstinence was associated with CHW visit number (OR=1.85, 95% CI=[1.29, 2.66]) and duration (OR=1.85, 95% CI=[1.33, 2.58]) and number of groups attended (OR=1.51, 95% CI=[1.00, 2.28]); effects on pharmacotherapy initiation were similar. 1-3 CHW visits per month over two years was optimal for achieving abstinence. Interviews identified engagement facilitators, including CHWs establishing trust, providing goal accountability, skills reinforcement, and assistance overcoming barriers to treatment access and adherence related to social determinants of health and illness factors. Robust training and supervision facilitated CHW effectiveness. Barriers included PCPs’ and care teams’ limited understanding of the CHW role.

**Conclusions:** Feasible CHW engagement was associated with tobacco abstinence in adults with serious mental illness. CHW implementation may benefit from promoting CHW training and integration within clinical teams.

**Highlights:**

- Greater participant engagement with community health workers (CHWs) (e.g., visit number, duration) was associated with higher tobacco abstinence rates in a two-year intervention for adults with serious mental illness and tobacco use disorder.
- Interviews with participants, CHWs, and primary care providers (PCPs) indicated that CHWs built trust, facilitated health behavior change, helped participants overcome adverse social determinants of health and other barriers to obtaining effective tobacco use disorder treatment.
- Training and supervision were perceived to be essential to CHW effectiveness; CHWs and PCPs suggested insufficient integration of CHWs into psychiatric rehabilitation and healthcare teams as an addressable implementation barrier.

## INTRODUCTION

High rates of tobacco use disorder and tobacco-related illness[1,2,3], drive premature mortality in those with serious mental illness[3]. Tobacco cessation pharmacotherapies, particularly varenicline, are effective and safe for this population[4–6], but underutilized[2,8,9], due to provider knowledge gaps[10,11] and low prioritization of tobacco cessation treatment in psychiatric care[9]. Further, those with serious mental illness have lower cessation treatment adherence[10], attributable to illness-related psychiatric and cognitive symptoms and to adverse social determinants of health that exacerbate barriers to treatment access and adherence[11].

Innovations are imperative to overcome multi-level barriers[12] and promote initiation and adherence to first-line tobacco use disorder healthcare to reduce tobacco-related morbidity and mortality in this population. Community health workers (CHWs) are frontline public health workers who can bridge treatment gaps for marginalized populations and improve hypertension, diabetes, and HIV/AIDS outcomes[13] by providing care coordination and assistance with treatment uptake and adherence[14] health education, advocacy, coaching/counseling, case management, and addressing health-related social needs[15]. In a recent pragmatic trial[16], we demonstrated that CHW support to adults with serious mental illness and tobacco use disorder and their primary care providers (PCPs) for two years, combined with provider education (PE), increased bio-verified tobacco abstinence rates over usual care or PE alone, an effect mediated through increased varenicline initiation and through an independent effect of CHW support. Participants who received tobacco cessation pharmacotherapy with CHW support were over three-fold more likely to quit smoking than those initiating pharmacotherapy without CHW support[16]. To guide CHW implementation efforts, we undertook a secondary analysis to identify effective dose ranges of CHW intervention components, understand components of the CHW intervention viewed as critical by patients, CHWs and PCPs[17], and identify barriers and facilitators of implementing the CHW intervention.

## METHODS

### Study Design

This study utilized a parallel mixed-methods design to determine effective type (one-to-one CHW visits, CHW co-led smoking cessation groups, CHW-accompanied PCP visits) and dose (number, duration) of participant-CHW engagement. To identify barriers and facilitators to implementing the CHW intervention, we conducted post-intervention interviews with stakeholders participating in the PE+CHW arm. The study was approved by Mass General Brigham and Massachusetts Department of Mental Health institutional review boards.

### CHW Intervention

In the parent trial (11/2017 to 01/2020)[16], primary care clinics were cluster-randomized to PE or usual care; PE-assigned clinics had a nested, participant-level randomization to be offered CHW support or not. Participants did not need to express willingness to quit smoking to enroll. The content and delivery of the CHW intervention was flexible, largely determined by individual participant characteristics and preferences. CHWs were twelve bachelor’s-level staff without clinical training or tobacco cessation experience who completed a two-week training covering core competencies for CHW certification (https://www.mass.gov/info-details/core-competencies-for-community-health-workers), agency-mandated safety training, Tobacco Treatment Specialist Core Training[18], skills to facilitate illness self-management, and education about the safety and efficacy of first-line tobacco cessation pharmacotherapies. CHWs visited participants in their homes or neighborhoods, co-led 60-minute-long, community-based smoking cessation group counselling sessions with a clinically-trained study staff member, encouraged participants to discuss their tobacco use, set smoking cessation goals, educated participants about safety and efficacy and encouraged trials of cessation medications, assisted participants at PCP visits and smoking cessation groups, aided communication with PCPs and community support staff to improve treatment adherence, and addressed participants’ unmet social determinants of health[16]. CHWs received weekly clinical group and as-needed individual supervision, and charted participant encounters.

#### Quantitative Method

##### Participants

Of 336 adults with tobacco use disorder and serious mental illness eligible for state psychiatric rehabilitation services assigned to the PE+CHW arm of the RCT, 116 (35%) did not consent to CHW support, and 21 (6%) consented participants dropped out before the Year 1 assessment. 196 retained participants were included in the analysis after excluding data from three outliers with ≥120 CHW visits (average SD above mean=4.2).

##### Data Collection and Measures

Data on participants-CHW engagement were extracted from CHW-reported weekly progress notes, consisting number/duration of in-person CHW visits, smoking cessation groups attended, and CHW-attended PCP visits and converted to z-scores for regression analyses. Outcomes were assessed blindly at Years 1 and 2 and included participant report of taking ≥1 dose of any tobacco cessation pharmacotherapy (nicotine replacement therapy, bupropion, varenicline) and abstinence (self-report of no tobacco smoking in the prior week and expired carbon monoxide of ≤5 parts/million [19]).

##### Statistical Analysis

Missing data for Year 2 tobacco abstinence in 43 participants were imputed ten times via predictive mean-matching using Year 1 abstinence outcomes and baseline characteristics, per the parent study [16]. We tested linear and quadratic relationships between each CHW engagement dose parameter, as a z-score, and each outcome using logistic regression. Inclusion of a quadratic term allowed testing of whether participants with the highest engagement may have diminishing returns[20]. We evaluated goodness-of-fit of model predictions by visually comparing predicted response proportions against observed proportions. For each outcome-engagement variable pair, observed percentage response and 95% CI were calculated within each bin of dose parameter. Bin sizes were determined by the Freedman-Diaconis algorithm[21]. To identify effective doses of CHW intervention components, we calculated the minimum threshold associated with significant improvement (i.e., expected response on outcome exceeds 95% prediction interval for the response given no intervention), and the maximum threshold with no further significant improvement (i.e., expected response falls below the 95% prediction interval for the highest possible predicted response)[22]. Analyses were conducted using R[23] with missing data imputed using the ‘mice’ package[24].

#### Qualitative Method

##### Participants

All twelve CHWs were interviewed. Study participants randomized to CHW support who completed the two-year intervention (N=153) were eligible for interview. All PCPs who received PE (N=111) were contacted for interview. Participant interviewees were selected using purposive sampling to represent a range of treatment outcomes achieved (tobacco abstinence, tobacco cessation pharmacotherapy initiated, percentage of behavioral smoking goals achieved in CHW sessions). Participants were interviewed until thematic saturation was reached. PCPs interviewed were a convenience sample based on interest in participating. Interviewees provided written informed consent and were compensated $15.

##### Interview Procedure

Semi-structured, individual interviews were conducted post-intervention to capture stakeholders’ experiences, attitudes, and perceived barriers and facilitators to effective engagement with and implementation of the CHW intervention. Interviews were conducted in English, in-person or via telephone/videoconference (MM, KS, DA), audio-recorded, and transcribed verbatim, except for one participant who refused audio-recording and detailed notes were used.

##### Qualitative Analysis

Team members (CF, LN, AR) independently coded transcribed interviews. We analyzed transcripts from each stakeholder group separately using a coding reliability approach to thematic analysis[25], first applying an *a priori*, deductive coding frame of higher-order domains based on the Consolidated Framework for Implementation Research (CFIR)[26], a framework designed to identify multilevel barriers and facilitators at the systemic, organizational, user, and intervention levels. Next, team members inductively identified themes that were confirmed by team discussion and consensus then mapped onto CFIR constructs[27]. Intercoder reliability was implemented in up to 25% of interviews from each stakeholder group until satisfactory reliability (κ ≥ 0.75) was achieved[28,29]. Coding differences and codebook refinements were resolved after each round of coding to ensure themes were comprehensively and accurately captured across all transcripts. We reviewed and synthesized all codes across stakeholder groups to identify major salient themes with consideration of lesser endorsed codes that were relevant and important to informing future implementation of the CHW intervention. Qualitative analysis was managed using NVivo 12 for Mac[30]. See online supplement for detailed analytic method.

## RESULTS

### Quantitative Results

#### Dose effects of CHW engagement

196 participants (30% female; 1% American Indian/Alaskan Native; 2% Asian; 39% Black/African American; 17% Hispanic/Latinx; 3% multiracial; 37% non-Hispanic white; moderate nicotine dependence on Heaviness of Smoking Index [31]; mean age: 49-years-old) were included in the analysis. Participants who consented and provided Year 1 data were more likely to be in supervised housing (51% vs. 32.1%, p=.012); enrollment and retention did not significantly differ by other characteristics (see online supplement).

Greater number and longer duration of CHW visits and more group visits attended were generally associated with higher likelihood of smoking cessation and tobacco cessation pharmacotherapy initiation (Figure 1). Participants were more likely to quit given a standard deviation unit increase in CHW visit number (OR=1.85, 95% CI=[1.29, 2.66]; SD=28 visits), CHW visit duration (OR=1.85, 95% CI=[1.33, 2.58]; SD=14 minutes), and number of smoking cessation groups attended (OR=1.51, 95% CI=[1.00, 2.28]; SD=23 sessions) (Table 1). Dose effects for tobacco cessation pharmacotherapy initiation were slightly larger and included a significant effect from CHW-attended PCP visit number (OR=5.64, 95% CI=[2.67, 11.91]; SD=3 visits).

**Figure 1:**
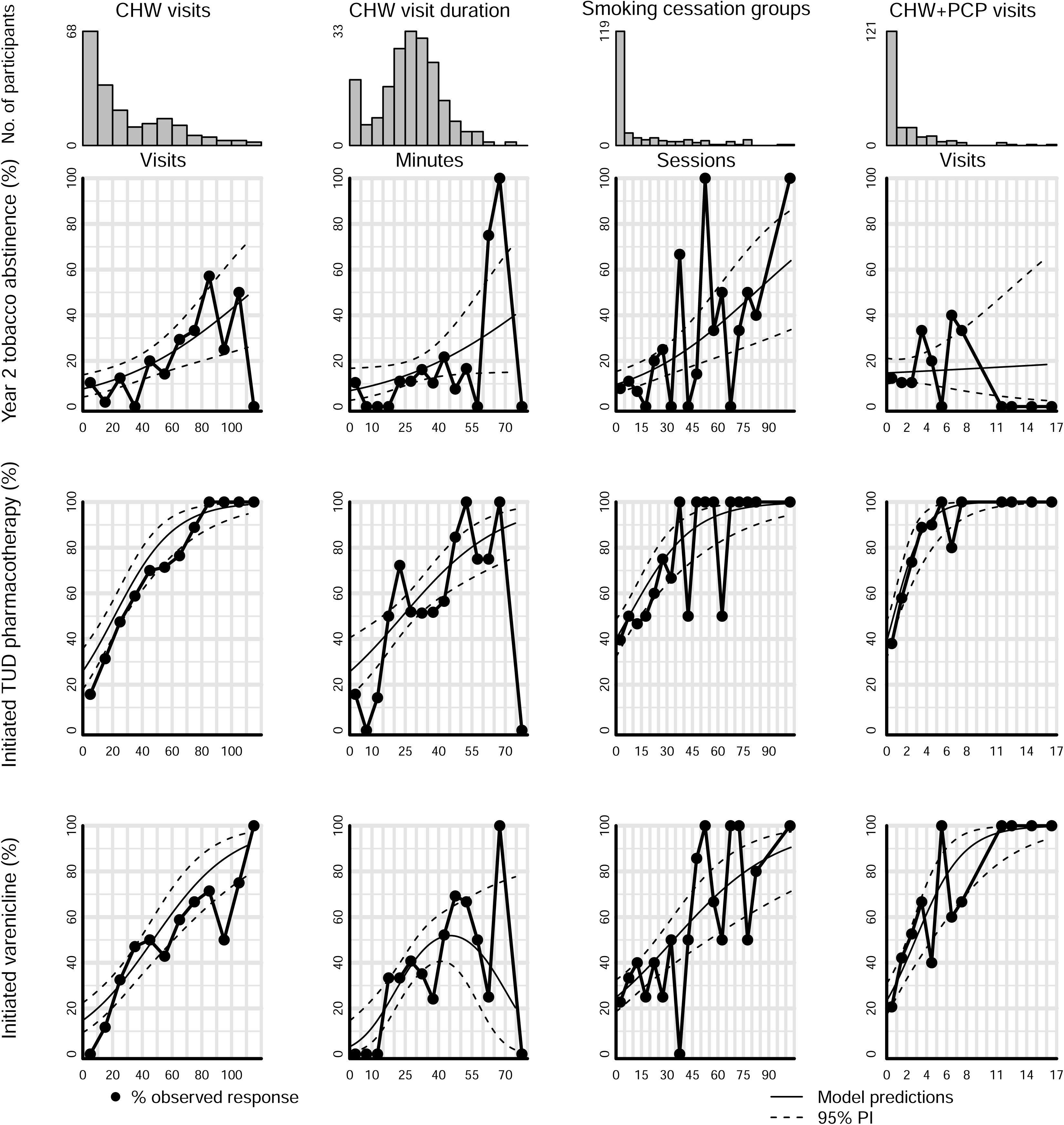
Observed and predicted dose-response. Plot of observed response (y-axis) by dose (binned over fixed intervals; x-axis) for each dose-response pair. Each column corresponds to an engagement variable; each row corresponds to a smoking cessation outcome variable. Observed percentage response and 95% confidence intervals (CI) based on a beta-binomial model were calculated over bin sizes determined by the Freedman-Diaconis algorithm [21]. Model predictions and 95% prediction intervals (PI) from a logistic regression model were overlaid. Models have good fit to the observed data, with most binned responses falling within 95% PI. Histograms presenting the observed data distribution from participants are shown at the top of each figure panel for each engagement variable.

**Table 1:** Summary of dose analysis by engagement variable and smoking cessation treatment outcome variable (N=196)

| Outcome/Engagement | Test statistics |  |  | Expected response with no Intervention | Thresholds for significant improvement |  | Expected responses at at min and max thresholds |  | Received ≥ min threshold |  | Received ≥ max threshold |  |
| --- | --- | --- | --- | --- | --- | --- | --- | --- | --- | --- | --- | --- |
|  | 95% CI |  |  |  | Min | Max | Min | Max | N | % | N | % |
|  | OR | LL | UL |  |  |  |  |  |  |  |  |  |
| Year 2 Tobacco Abstinence |  |  |  |  |  |  |  |  |  |  |  |  |
| CHW visits | 1.85* | 1.29 | 2.66 | 7.7 | 30 | 65 | 14.0 | 25.9 | 71 | 36 | 26 | 13 |
| CHW visit duration <sup>a</sup> | 1.51* | 1.00 | 2.28 | 7.0 | 29 | 34 | 15.1 | 16.8 | 92 | 47 | 58 | 30 |
| Smoking cessation groups | 1.85* | 1.33 | 2.58 | 9.7 | 20 | 57 | 15.4 | 33.5 | 50 | 26 | 14 | 7 |
| CHW-attended PCP visits <sup>b</sup> | 1.05 | 0.72 | 1.52 | 14.7 | N/A | N/A | N/A | N/A | N/A | N/A | N/A | N/A |
| Initiated TUD pharmacotherapy |  |  |  |  |  |  |  |  |  |  |  |  |
| CHW visits | 4.03* | 2.53 | 6.44 | 25.9 | 9 | 79 | 35.8 | 94.6 | 129 | 66 | 13 | 7 |
| CHW visit duration | 1.88* | 1.36 | 2.59 | 25.7 | 15 | 49 | 40.7 | 75.7 | 163 | 83 | 10 | 5 |
| Smoking cessation groups | 3.32* | 1.97 | 5.61 | 39.9 | 7 | 61 | 48.7 | 94.3 | 72 | 37 | 13 | 7 |
| CHW-attended PCP visits | 5.64* | 2.67 | 11.91 | 39.0 | 1 | 9 | 48.1 | 99.6 | 75 | 38 | 6 | 3 |
| Initiated Varenicline |  |  |  |  |  |  |  |  |  |  |  |  |
| CHW visits | 2.84* | 1.98 | 4.08 | 14.8 | 14 | 81 | 22.5 | 78.1 | 117 | 60 | 12 | 6 |
| CHW visit duration <sup>c</sup> | 0.72* | 0.54 | 0.95 | 3.2 | 12 | 75 | 14.6 | 20.3 | 167 | 85 | N/A | N/A |
| Smoking cessation groups | 2.09* | 1.50 | 2.92 | 24.8 | 12 | 62 | 32.8 | 71.2 | 61 | 31 | 13 | 7 |
| CHW-attended PCP visits | 2.97* | 1.85 | 4.77 | 23.4 | 1 | 9 | 31.2 | 93.9 | 75 | 38 | 6 | 3 |

**Note.** OR: Odds ratio per one standard deviation increase in engagement variable units; CI: Confidence interval; LL: Lower limit; UL: Upper limit; N: Number of participants; CHW: community health worker; TUD: tobacco use disorder; PCP: primary care provider. \* p < .05.
<sup>a</sup> Limits of prediction intervals for lowest and highest predicted response overlap.
<sup>b</sup> No significant dose-response relationship.
<sup>c</sup> Odds ratio reported for significant quadratic trend for dose-response relationship. Linear OR: 2.37 [1.51, 3.74], p < .001

The optimal dosage ranges of CHW support for Year 2 tobacco abstinence were 30-65 visits over two years (e.g., 1-3 visits per month), 29-34 minutes per visit (e.g., half-hour visits), and 20-57 CHW co-led smoking cessation groups (e.g., 1-3 groups per month) (Table 1). Increasing the dose of CHW engagement above these levels led to only marginal improvements in tobacco cessations. For CHW visits, visit duration, and cessation groups, 36%, 47%, and 26% of participants, respectively, received this threshold dose.

### Qualitative Results

Twelve CHWs (83% female; 67% non-Hispanic white; 25% Hispanic/Latinx; 8% Black; mean age: 27-years-old), 16 participants (Table 2) and 17 PCPs (47% female; 94% physicians; 6% nurse practitioners) were interviewed. Generally, all groups valued all types of CHW intervention (CHW visits, smoking cessation groups, and CHW-attended PCP visits) and perceived them to be integral to participants’ tobacco reduction or cessation.

**Table 2:** Characteristics of interviewed patient participants (N=16)

| Measure | % | M±SD/N |
| --- | --- | --- |
| ***Demographic characteristics at baseline*** |  |  |
| Age (years) |  | 49.5±11.2 |
| Sex (female) | 29.4 | 5 |
| Race |  |  |
| American Indian or Alaskan Native <sup>a</sup> | 0 | 0 |
| Asian <sup>a</sup> | 5.9 | 1 |
| Black or African American | 52.9 | 9 |
| Hispanic or Latino | 23.5 | 4 |
| Multiracial | 0 | 0 |
| Non-Hispanic white | 17.6 | 3 |
| Supervised housing | 35.3 | 6 |
| SF-1 <sup>b</sup> |  | 2.9±1.0 |
| Expired CO (ppm) <sup>c</sup> |  | 22.0±20.3 |
| HIS <sup>d</sup> |  | 2.8±1.5 |
| Tobacco products per day |  | 11.3±7.4 |
| Cigarettes (daily use) | 88.2 | 15 |
| Little cigars (daily use) | 47.1 | 8 |
| Hand-rolled cigarettes (daily use) | 5.9 | 1 |
| E-cigarettes (daily use) | 0 | 0 |
| Physician recommendation to quit smoking | 58.8 | 10 |
| Prior year prescribed medication to aid cessation | 41.2 | 7 |
| Cardiovascular/respiratory illness | 41.2 | 7 |
| Other smoking related illness <sup>e</sup> | 35.3 | 6 |
| ***Treatment and engagement outcomes over two years*** |  |  |
| Year 2 tobacco abstinence | 17.6 | 3 |
| Initiated TUD pharmacotherapy | 76.5 | 13 |
| Initiated varenicline | 52.9 | 9 |
| Quantity of CHW visits |  | 40.8±32.1 |
| Average duration of CHW visits |  | 31.5±11.6 |
| Quantity of smoking cessation groups |  | 27.8±31.9 |
| Quantity of CHW-attended PCP visits |  | 2.0±4.0 |

**Note.** M: Mean; SD: Standard deviation; N: Number of participants; TUD: tobacco use disorder; CHW: community health worker; PCP: primary care provider.
<sup>a</sup>. Insufficient cell count (≤4 participants) for analyses
<sup>b</sup>. SF-1 is single-item self-report of overall health. Possible scores range from 1 to 5, corresponding to perceived health being poor, fair, good, very good, or excellent.
<sup>c</sup>. Expired air carbon monoxide (CO) is a measure of current smoking; mean CO reported here is consistent with smoking approximately 1 pack per day. Abstinence was defined as ≤ 5 parts per million (ppm) of CO.
<sup>d</sup>. Possible scores on the Heaviness of Smoking Index (HSI) range from 0 to 6; scores of 0–2 indicate low severity of nicotine dependence, 3 and 4 indicate moderate dependence, and 5 and 6 indicate severe dependence.
<sup>e</sup>. Includes diabetes, cancer, pneumonia, tuberculosis, cataracts, glaucoma, and retinal disease

We found five key barriers and facilitators to effective implementation of CHW support specifically, categorized into three major themes, presented in Table 3 along with their corresponding CFIR domains, illustrative quotes, and implementation recommendations. See online supplement for implementation factors for PE and smoking cessation groups.

**Table 3:**
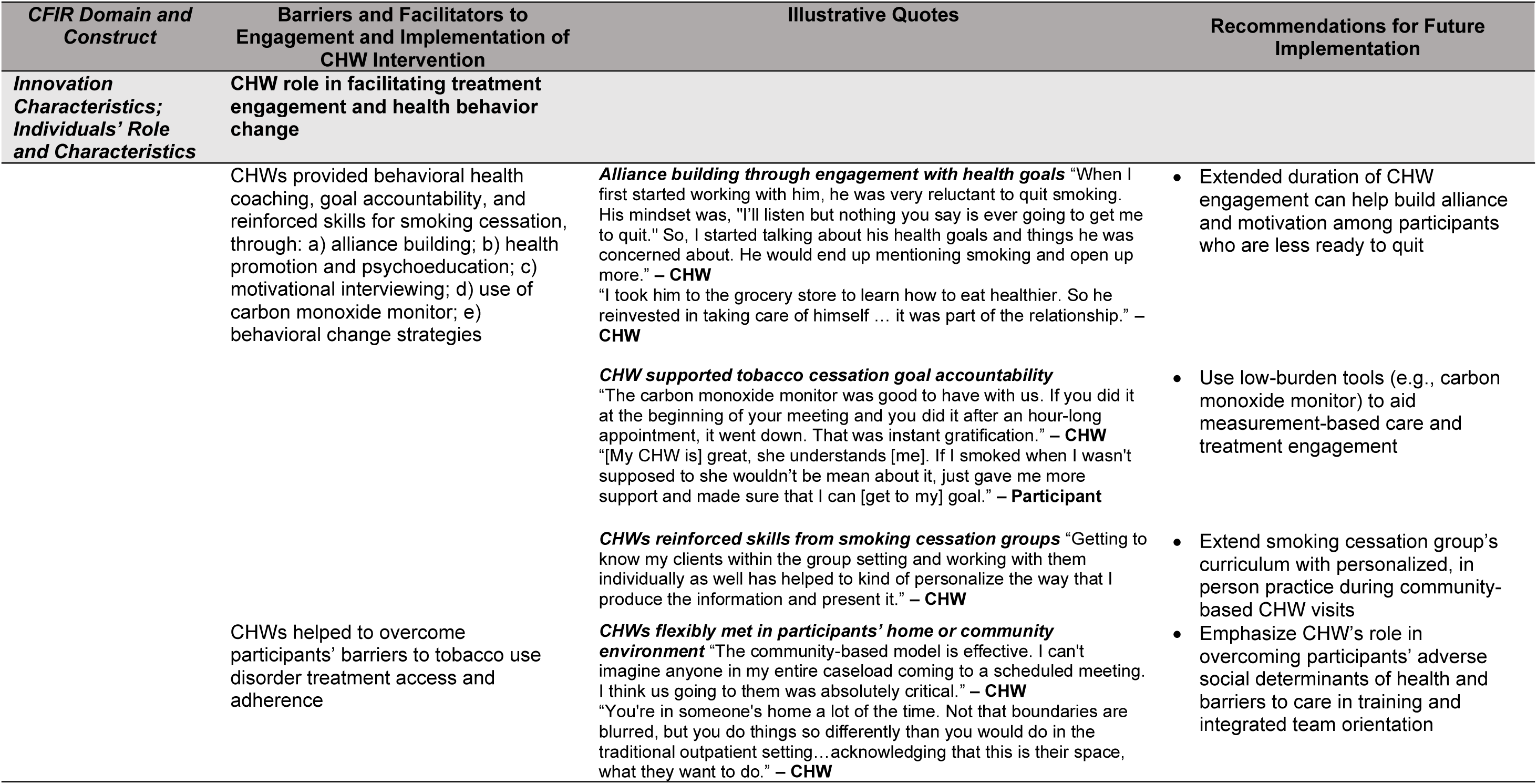
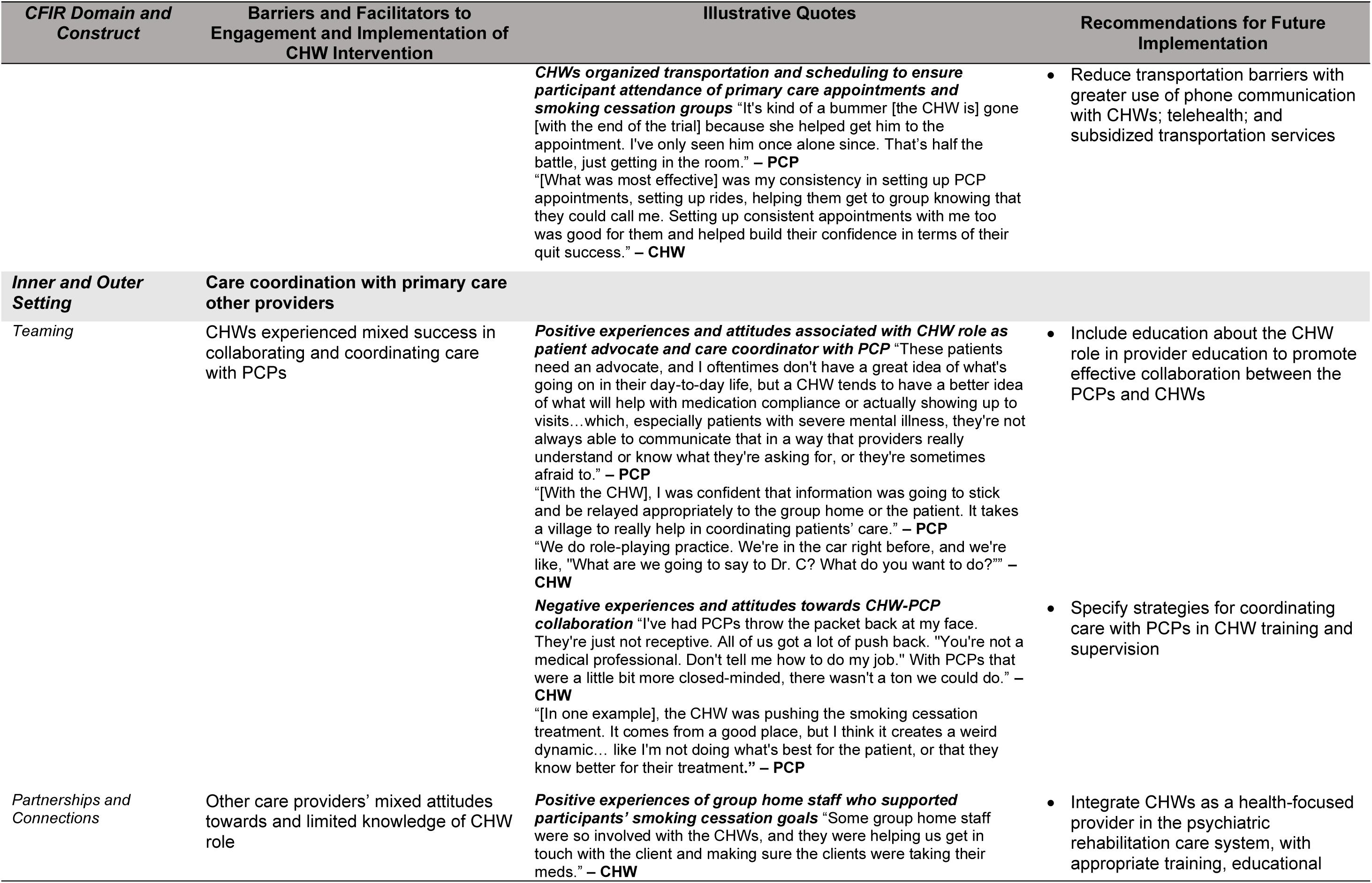
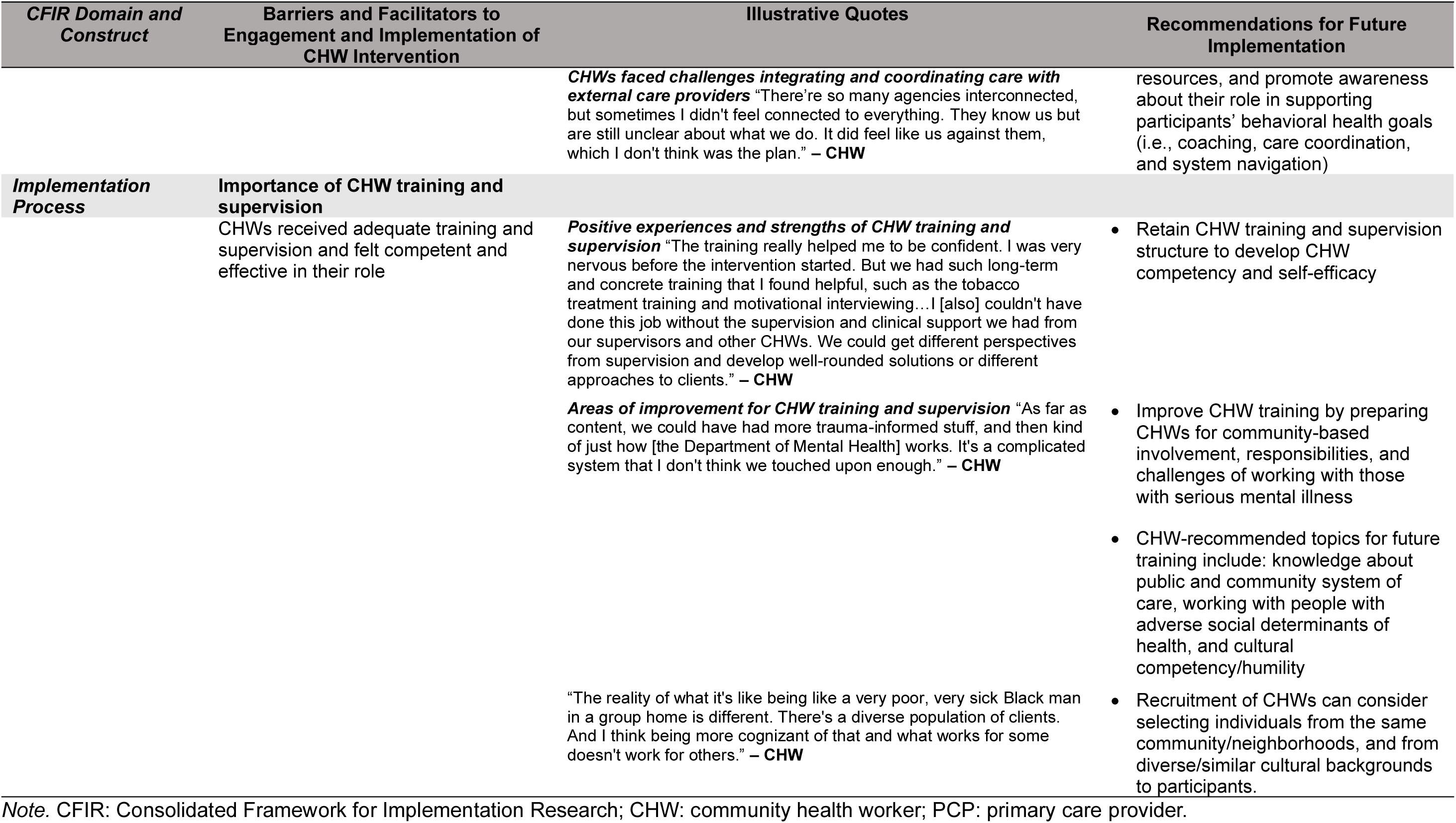
CHW, PCP, and study participants’ perceived barriers and facilitators to engagement with and implementation of CHW intervention.

| *CFIR Domain and Construct* | **Barriers and Facilitators to Engagement and Implementation of CHW Intervention** | **Illustrative Quotes** | **Recommendations for Future Implementation** |
| --- | --- | --- | --- |
| ***Innovation Characteristics; Individuals' Role and Characteristics*** | **CHW role in facilitating treatment engagement and health behavior change**<br>CHWs provided behavioral health coaching, goal accountability, and reinforced skills for smoking cessation, through: a) alliance building; b) health promotion and psychoeducation; c) motivational interviewing; d) use of carbon monoxide monitor; e) behavioral change strategies<br>CHWs helped to overcome participants' barriers to tobacco use disorder treatment access and adherence | ***Alliance building through engagement with health goals*** "When I first started working with him, he was very reluctant to quit smoking. His mindset was, "I'll listen but nothing you say is ever going to get me to quit." So, I started talking about his health goals and things he was concerned about. He would end up mentioning smoking and open up more." – **CHW**<br>"I took him to the grocery store to learn how to eat healthier. So he reinvested in taking care of himself ... it was part of the relationship." – **CHW**<br>***CHW supported tobacco cessation goal accountability***<br>"The carbon monoxide monitor was good to have with us. If you did it at the beginning of your meeting and you did it after an hour-long appointment, it went down. That was instant gratification." – **CHW**<br>"[My CHW is] great, she understands [me]. If I smoked when I wasn't supposed to she wouldn't be mean about it, just gave me more support and made sure that I can [get to my] goal." – **Participant**<br>***CHWs reinforced skills from smoking cessation groups*** "Getting to know my clients within the group setting and working with them individually as well has helped to kind of personalize the way that I produce the information and present it." – **CHW**<br>***CHWs flexibly met in participants' home or community environment*** "The community-based model is effective. I can't imagine anyone in my entire caseload coming to a scheduled meeting. I think us going to them was absolutely critical." – **CHW**<br>"You're in someone's home a lot of the time. Not that boundaries are blurred, but you do things so differently than you would do in the traditional outpatient setting...acknowledging that this is their space, what they want to do." – **CHW** | Extended duration of CHW engagement can help build alliance and motivation among participants who are less ready to quit<br>Use low-burden tools (e.g., carbon monoxide monitor) to aid measurement-based care and treatment engagement<br>Extend smoking cessation group's curriculum with personalized, in person practice during community-based CHW visits<br>Emphasize CHW's role in overcoming participants' adverse social determinants of health and barriers to care in training and integrated team orientation |

| **CFIR Domain and Construct** | **Barriers and Facilitators to Engagement and Implementation of CHW Intervention** | **Illustrative Quotes** | **Recommendations for Future Implementation** |
| --- | --- | --- | --- |
|  |  | **CHWs organized transportation and scheduling to ensure participant attendance of primary care appointments and smoking cessation groups** "It's kind of a bummer [the CHW is] gone [with the end of the trial] because she helped get him to the appointment. I've only seen him once alone since. That's half the battle, just getting in the room." – PCP<br>"[What was most effective] was my consistency in setting up PCP appointments, setting up rides, helping them get to group knowing that they could call me. Setting up consistent appointments with me too was good for them and helped build their confidence in terms of their quit success." – CHW | • Reduce transportation barriers with greater use of phone communication with CHWs; telehealth; and subsidized transportation services |
| **Inner and Outer Setting** | **Care coordination with primary care other providers** |  |  |
| *Teaming* | CHWs experienced mixed success in collaborating and coordinating care with PCPs | **Positive experiences and attitudes associated with CHW role as patient advocate and care coordinator with PCP** "These patients need an advocate, and I oftentimes don't have a great idea of what's going on in their day-to-day life, but a CHW tends to have a better idea of what will help with medication compliance or actually showing up to visits...which, especially patients with severe mental illness, they're not always able to communicate that in a way that providers really understand or know what they're asking for, or they're sometimes afraid to." – PCP<br>"[With the CHW], I was confident that information was going to stick and be relayed appropriately to the group home or the patient. It takes a village to really help in coordinating patients' care." – PCP<br>"We do role-playing practice. We're in the car right before, and we're like, "What are we going to say to Dr. C? What do you want to do?" – CHW<br>**Negative experiences and attitudes towards CHW-PCP collaboration** "I've had PCPs throw the packet back at my face. They're just not receptive. All of us got a lot of push back. "You're not a medical professional. Don't tell me how to do my job." With PCPs that were a little bit more closed-minded, there wasn't a ton we could do." – CHW<br>"[In one example], the CHW was pushing the smoking cessation treatment. It comes from a good place, but I think it creates a weird dynamic... like I'm not doing what's best for the patient, or that they know better for their treatment." – PCP | • Include education about the CHW role in provider education to promote effective collaboration between the PCPs and CHWs<br>• Specify strategies for coordinating care with PCPs in CHW training and supervision |
| *Partnerships and Connections* | Other care providers' mixed attitudes towards and limited knowledge of CHW role | **Positive experiences of group home staff who supported participants' smoking cessation goals** "Some group home staff were so involved with the CHWs, and they were helping us get in touch with the client and making sure the clients were taking their meds." – CHW | • Integrate CHWs as a health-focused provider in the psychiatric rehabilitation care system, with appropriate training, educational |
|  |  | **CHWs faced challenges integrating and coordinating care with external care providers** "There're so many agencies interconnected, but sometimes I didn't feel connected to everything. They know us but are still unclear about what we do. It did feel like us against them, which I don't think was the plan." – CHW | resources, and promote awareness about their role in supporting participants' behavioral health goals (i.e., coaching, care coordination, and system navigation) |
| **Implementation Process** | **Importance of CHW training and supervision**<br>CHWs received adequate training and supervision and felt competent and effective in their role | **Positive experiences and strengths of CHW training and supervision** "The training really helped me to be confident. I was very nervous before the intervention started. But we had such long-term and concrete training that I found helpful, such as the tobacco treatment training and motivational interviewing...I [also] couldn't have done this job without the supervision and clinical support we had from our supervisors and other CHWs. We could get different perspectives from supervision and develop well-rounded solutions or different approaches to clients." – CHW<br>**Areas of improvement for CHW training and supervision** "As far as content, we could have had more trauma-informed stuff, and then kind of just how [the Department of Mental Health] works. It's a complicated system that I don't think we touched upon enough." – CHW<br>"The reality of what it's like being like a very poor, very sick Black man in a group home is different. There's a diverse population of clients. And I think being more cognizant of that and what works for some doesn't work for others." – CHW | • Retain CHW training and supervision structure to develop CHW competency and self-efficacy<br>• Improve CHW training by preparing CHWs for community-based involvement, responsibilities, and challenges of working with those with serious mental illness<br>• CHW-recommended topics for future training include: knowledge about public and community system of care, working with people with adverse social determinants of health, and cultural competency/humility<br>• Recruitment of CHWs can consider selecting individuals from the same community/neighborhoods, and from diverse/similar cultural backgrounds to participants. |

*Note.* CFIR: Consolidated Framework for Implementation Research; CHW: community health worker; PCP: primary care provider.

#### Facilitating treatment engagement and behavioral change

Endorsed by all stakeholder groups, this theme relates to the innovation characteristics CFIR domain, elaborating on the critical functions of the CHW role. Participants emphasized the importance of building trust and CHWs stressed the importance of alliance building, focusing on general health goals for those not ready to quit smoking. CHWs and participants reported that CHWs facilitated goal accountability by educating on evidence-based smoking cessation practices, using motivational interviewing, and reinforcing health behavior change by assisting participants to practice skills learned from groups (e.g., identifying triggers and implementing coping skills) in their neighborhoods. Using carbon monoxide monitoring during CHW visits helped quantify participants’ smoking reduction or abstinence, which was reported to facilitate CHWs’ ability to reinforce participant progress and problem-solve obstacles to meeting smoking cessation goals, thus enhancing participant engagement and CHWs’ sense of competence.

All stakeholders highlighted how CHWs facilitated participant engagement by helping participants overcome barriers to treatment access and adherence (e.g., scheduling and organizing transportation to PCP appointments and groups; navigating insurance lapses for maintenance of pharmacotherapy; collaborating to devise workable medication adherence regimens), address adverse social determinants of health (e.g., reducing transportation barriers by meeting participants in their homes/neighborhood; providing transport to grocery shopping; reducing social isolation), and mitigate the impact of cognitive and motivational deficits associated with serious mental illness and shortfalls in health/mental health literacy.

#### Care coordination challenges with primary care and other providers

This theme highlighted mixed success in effective teaming within primary care (inner setting) and building partnerships with the psychiatric rehabilitation team (outer setting). Most interviewees reported positive experiences of CHWs assisting participants in advocating for their health goals with PCPs, obtaining prescriptions for tobacco cessation pharmacotherapy, and liaising with participants’ psychiatric rehabilitation teams to support medication adherence. However, some CHWs encountered resistance from PCPs with respect to their role on the participants’ care teams, describing instances where their input on cessation medication use was dismissed. A few PCPs expressed concerns about CHWs potentially undermining their clinical authority and decisions.

CHWs also had mixed experiences coordinating care with other psychiatric rehabilitation providers. For example, some CHWs reported that residential staff had little understanding of the CHW role, had conflicting approaches to smoking cessation (e.g., using punitive measures, or providing cigarettes as a reward), and were unresponsive to care coordination efforts.

#### Importance of CHW training and supervision

Critical to the implementation process, nearly all CHWs spoke positively about pre-intervention specialized training and ongoing clinical supervision, which helped them handle challenging clinical situations and develop competency. Many found value in the apprenticeship learning model, involving shadowing clinical outreach teams before taking cases. See Table 3 for CHW recommendations for future trainings.

## DISCUSSION

Greater engagement between participants and CHWs was associated with higher tobacco abstinence rates in adults with serious mental illness. Increased tobacco abstinence rates were observed with highly feasible CHW intervention dose ranges of 1-3 30-minute CHW visits and 1-3 CHW co-led smoking cessation groups per month over two years. These doses were received by over a quarter of participants. Study participants, CHWs, and PCPs identified factors influencing successful CHW intervention implementation, and specific CHW critical functions that helped overcome participants’ barriers to tobacco cessation treatment uptake and adherence in a primary care context.

Ideally, implementation of an evidence-based CHW intervention must strike a balance between the maximally efficacious dose and the dose that is reasonably affordable and feasibly accepted by a large enough proportion of recipients to be beneficial in real-world settings. As tobacco use disorder is a chronic condition and study participants were not necessarily interested in quitting smoking at enrollment, it was not surprising that an extended duration intervention was associated with better tobacco cessation outcomes[32,33]. We have shown previously that when offered extended duration tobacco cessation treatment, tobacco abstinence rates increase over time in people with serious mental illness, with the greatest increase in abstinence occurring after one year of treatment in those not ready to quit right away[34], replicating prior work with people with depressive disorders who smoked[35]. Central to our approach was an opt-out approach by which CHWs recommended tobacco cessation pharmacotherapy initiation in all participants, even those not immediately ready to quit[36], an approach now incorporated in clinical best practices[37]. We suspect tobacco cessation pharmacotherapy use was important for enhancing motivation and self-efficacy for cessation via mechanisms of extinction, reduced enjoyment of tobacco, and reduced craving. Consistent engagement over time in participants’ homes and communities was essential for building trust with participants, particularly those not initially interested in quitting, by engaging and motivating participants first through discussing general health goals and providing practical assistance in addressing barriers related to social determinants of health. Multiple quit attempts during the intervention may have led to more frequent CHW engagement, such that future testing of a bidirectional relationship between CHW engagement and outcomes is warranted. Future implementation study may also explore CHW extension through telehealth[38], mobile apps[39], and group formats to promote scalability of the CHW intervention.

Another key finding was the identification of CHW-attended PCP visits as a high-yield strategy for increasing initiation of tobacco cessation pharmacotherapy. Nearly half of participants who had a single CHW-attended PCP visit used tobacco cessation pharmacotherapy, and almost 25% used varenicline. This is an important outcome given the persistent low prescribing rates of nicotine replacement therapy (1.6%) and varenicline (2.4%) for individuals with serious mental illness and tobacco use disorder[7]. CHWs overccame barriers to physician prescribing by addressing transportation challenges to PCP visits; advocating for participants’ desire for treatment and reinforcing provider education about safety and efficacy of first-line tobacco cessation pharmacotherapy in people with serious mental illness during PCP visits; serving as a communication liaison with PCPs around adverse effects; navigating insurance lapses that interrupted prescription access; and communicating with participants’ psychiatric rehabilitation team to support medication adherence.

Concordant with trials demonstrating improved health behaviors impacting chronic health conditions in primary care settings with CHW coaching[40,41], qualitative results highlighted that CHWs amplified the benefits of skills-based smoking cessation groups and facilitated treatment engagement by overcoming barriers due to adverse social determinants of health, disease-related factors, cultural and linguistic barriers, and health system complexity[11,12,42].

Finally, multiple stakeholders emphasized the need for thoughtful integration of the CHW role within the healthcare system, such as a health-focused psychiatric rehabilitation worker who can support illness management across different care settings for people with serious mental illness. This finding is consistent with studies highlighting that coordination and collaboration of CHWs with other healthcare providers is critical for sustained implementation of CHW interventions[43,44]. Education about CHW effectiveness, propelled by recent calls for CHW professionalization[14,15] and recognition of their contributions to equitable provision of evidence-based care (e.g., in the Affordable Care Act,[45,46]), complemented with robust training and supervision, could support smoking cessation and potentially improve care for other chronic medical conditions in people with serious mental illness in community settings.

### Limitations

This analysis of participants with serious mental illness in an urban settings may have limited generalizability to individuals with less severe functional impairment, those not connected to primary care, other health systems or rural settings. Our dose-response analyses were based on observed doses conditional on participants’ consent to CHW support, limiting extrapolation outside the current observed data range and to those who may need longer periods of engagement to accept the CHW intervention. Future study of fixed target doses is warranted to test the dose range identified here. Dosing parameters may be confounded with intervention content (e.g., longer sessions may incorporate more skills)[47], although we assume fidelity to intervention due to robust CHW training and supervisory structure within the trial. We ensured trustworthiness of qualitative results by using reliable coding techniques, illustrative quotes, and external audit. However, PCP and patient participants who self-selected to interview were likely to be more engaged in the study, and perspectives from disengaged or non-consenting participants and administrator and policy-maker insights into system-level constraints and facilitators (e.g., capacity, budget) were not obtained.

## CONCLUSIONS

Greater CHW-participant engagement within feasible dose ranges was associated with higher tobacco cessation rates in adults with serious mental illness. Integrating CHWs into psychiatric rehabilitation teams and recognizing their role as behavioral health coaches, patient advocates, and care coordinators were perceived as important for successful implementation. Future research optimizing scalable delivery formats and cost-effectiveness for CHW interventions enhancing implementation and effectiveness of tobacco cessation treatment in psychiatric rehabilitation and primary care is warranted. There is potential to extend this effective behavioral health model to other health needs of those with serious mental illness, and other high-need populations facing treatment barriers related to social determinants of health.

## Data Availability

The complete de-identified dataset containing individual-participant level data that underpin the results reported in this article, together with the full study protocol, statistical analysis plan and statistical code used to generate the results will be archived, searchable, and open access, posted through the PCORI Patient-Centered Outcomes Data Repository, hosted by the University of Michigan's Inter-University Consortium for Political and Social Research (ICPSR. ICPSR is an open access repository that connects researchers to data funded by PCORI. According to PCORI's policy, the data requestor will enter a DUA and submit a request outlining their research purpose directly with the data repository. Their qualifications, research justification, data security plan, and assurance that the research will result in a generalizable, patient-relevant contribution will all be taken into account by a PCORI-designated review committee. Such requests and derived results are posted publicly. The DUA prohibits reidentification.

## Acknowledgements

The authors thank all participants and staff involved in the trial; the late Michael D. Fetters, MD, for his expert guidance on conceptualization of the mixed methods design; and Diana Arntz, for supporting qualitative data collection.

## ONLINE SUPPLEMENT

### Detailed Qualitative Analysis Methods

We include this supplement to provide detailed and transparent reporting of qualitative methods and analytic procedures. In the qualitative phase, we identified barriers and facilitators to engagement with and implementation of the community health worker (CHW)-supported tobacco cessation intervention from CHW, patient participant, and primary care physicians (PCP) who completed the two-year intervention.

Thematic analysis was chosen as the overarching qualitative analysis method as it is a systematic and flexible approach that aligns with the mixed methods objectives to complement the quantitative results, allowing for the deductive coding frame to be derived from theory-driven quantitative variables while still allowing for themes to emerge inductively from the data.^1,2^ Specifically, we undertook an inter-coder reliability (ICR) approach to thematic analysis^3^ to prioritize reliability of data coding, where themes were identified based on the agreement among multiple coders. An ICR procedure does not conflict with the interpretative goals of qualitative research; instead, it improves the systematicity and transparency of the coding process and demonstrates that a team of researchers working within a shared conceptual framework can arrive at a consensual understanding of the data. We analyzed transcripts from each stakeholder group separately (i.e., CHW, participant, PCP) using the ICR procedure illustrated in Figure A. Qualitative analysis was managed using NVivo 12 for Mac.

In Stage 1, CF developed an initial, deductive coding frame based on higher-order domains and sub-constructs from the Consolidated Framework for Implementation Research (CFIR),^4^ including: intervention characteristics of provider education, CHW support, and smoking cessation groups; outer setting; inner setting of primary care centers; and individual characteristics of CHWs and patient participants*. Intervention characteristics* included constructs related to perceived evidence strength and quality of the intervention, perceived difficulty of implementing the intervention, and perceived flexibility of the intervention to be adapted. *Outer setting* referred to external policies, incentives, other care entities, and structural barriers and facilitators that could influence the implementation of the intervention. *Inner setting* referred to characteristics and implementation readiness of primary care centers and primary care physicians, including structural characteristics, capacity and resources, values and behavioral norms of the organization, openness to and support of the intervention, and capacity of change. *Characteristics of CHWs* consisted of CHW attitudes towards the intervention, and beliefs about one’s own capabilities related to the implementation of the intervention. *Characteristics of patient participants* consisted of participants attitudes towards the intervention and other characteristics that may influence implementation of the intervention (e.g., individual stage of change, cognitive impairments, severity of illness). We included two higher-order categories related to the qualitative research question to assess successful CHW-support strategies and specific recommendations for future implementation.

Three team members trained in qualitative analysis and ICR (CF, LN, AR) independently coded transcripts by segmenting and labeling the text according to specific points that were raised. Themes were inductively developed by aggregating similar codes together, confirmed by team discussion and consensus, and then mapped onto the CFIR deductive frame. Constructs that did not have any relevant codes were removed. The output was an initial codebook that comprehensively, exhaustively, and parsimoniously captured codes across all transcripts from a stakeholder group, and organized into higher-order code categories based on the CFIR.^5^

In Stage 2, a stepwise method of ICR assessment was conducted where three to four randomly selected transcripts per stakeholder group (25% of sample) were double-coded to improve the reliability and transparency of the coding frame such that those using it would consistently apply the same codes to the same excerpts.^3^ Stepwise rounds of ICR testing were implemented until satisfactory reliability was achieved, with the coding frame refined after each round.^6,7^ The second coder received an uncoded file that highlights the segmented data units, but not their associated codes. The second coder then used the initial codebook to independently code the data units that were visible on the cleaned file. The first and second coder’s codes were compared, and reliability was calculated based on the Cohen’s kappa coefficient^8^ computed by NVivo 12, a statistical measure of ICR that corrects for agreement that could occur through chance, for the overall coding frame as well as individual codes. After comparison of one double-coded transcript, discussion meetings were held to expose similarities and differences in ways of applying codes and come to consensus in coding.^6,9^ Codes were more carefully described and operationalized during discussions between team members to produce a revised codebook for the next round of ICR until overall kappa coefficient of ≥ 0.75 for acceptable reliability was reached.^10^ In our study, we reached satisfactory reliability typically by two rounds of ICR for all three stakeholder groups.

In Stage 3, the finalized coding frame for each stakeholder group was applied to the remaining single-coded transcripts. We reviewed and synthesized all codes across stakeholder groups to identify major salient themes with consideration of lesser endorsed codes that were still relevant and important to informing future implementation of the CHW intervention. We reported our qualitative methods and results based on the Standards for Reporting Qualitative Research (SRQR) guideline to ensure transparency and rigor.^11^ Trustworthiness of our findings was strengthened by inter-coder reliability; rich and thick description of the cases with illustrative quotes reported as evidence; reviewing and reporting lesser endorsed and contrary evidence; subject matter expert external audit (AEE, AT, CC).

**Figure A.**
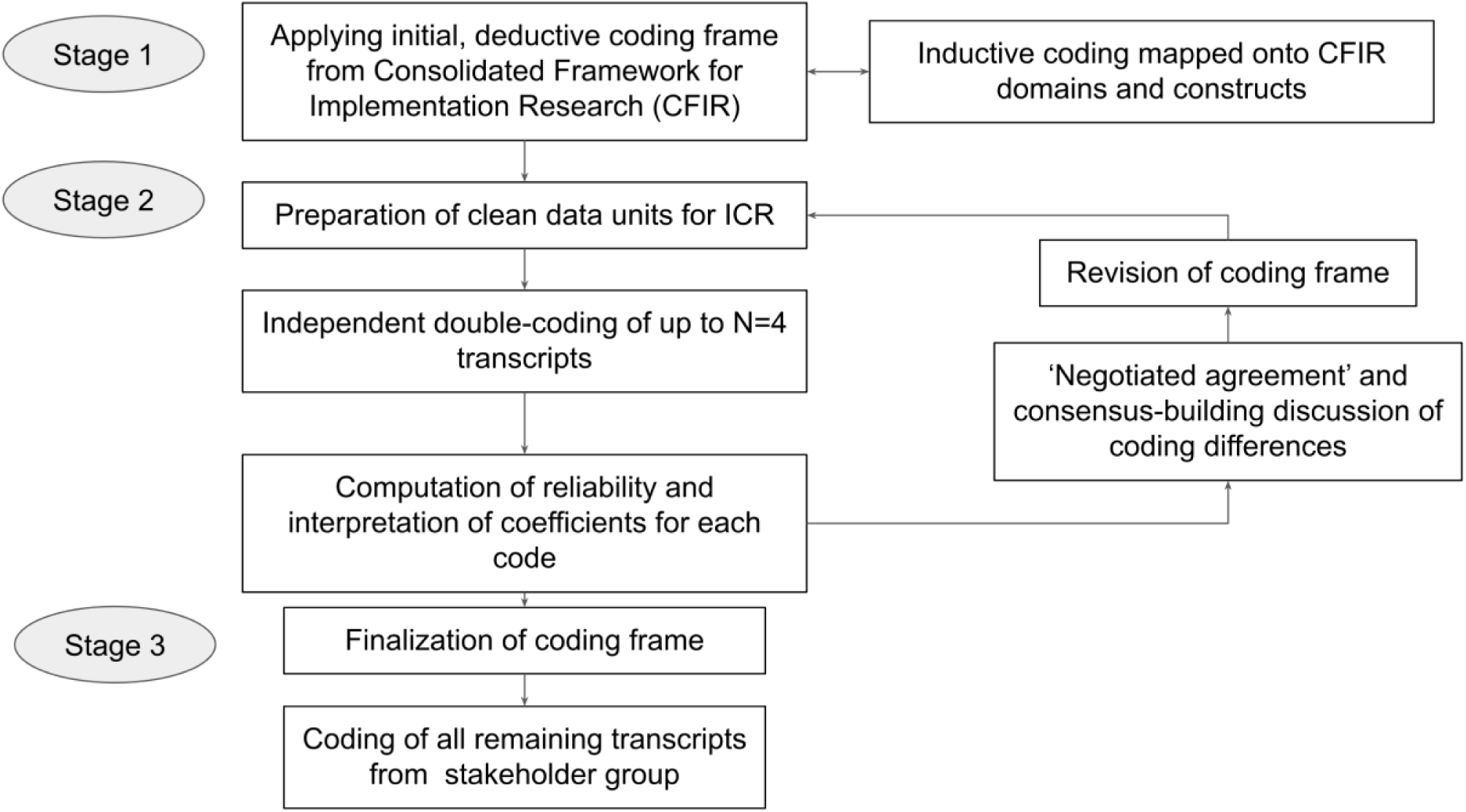
Inter-coder reliability procedure.

## Baseline characteristics of study participants by enrollment and retention

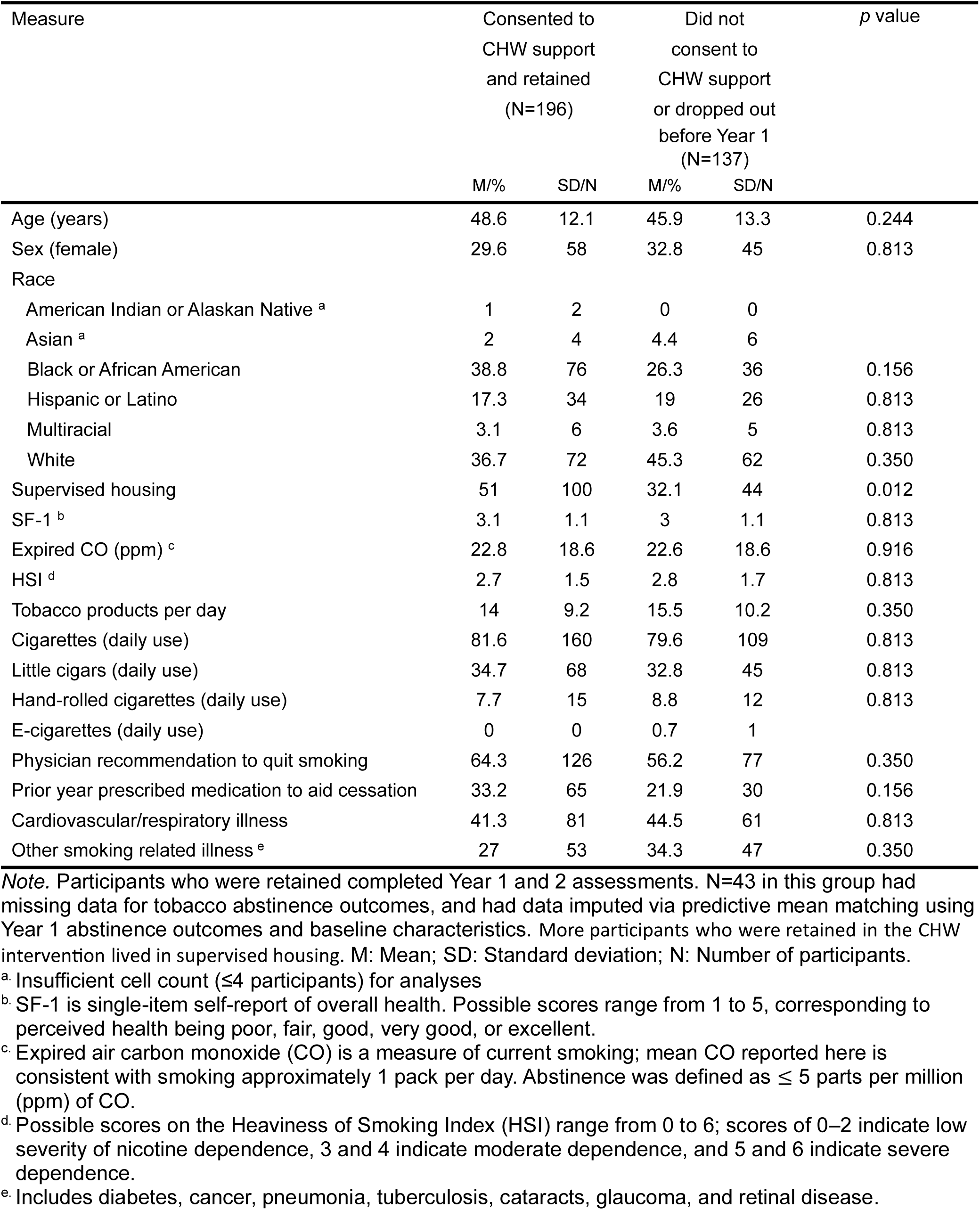

## Barriers and Facilitators to Engagement in Provider Education and Smoking Cessation Groups

In the parent clinical trial’s intervention group, primary care clinics received a one-time, clinician-delivered provider education (PE) on safety, efficacy, and importance of tobacco cessation pharmacotherapy; study participants who received care in PE-assigned clinics and were randomized to the intervention group also received up to two years of community health worker (CHW) support and weekly group counseling sessions on smoking cessation. Our primary focus was understanding facilitators and barriers to CHW engagement, but we were also interested in understanding the acceptability, perceived effectiveness, and factors influencing implementation for provider education (PE) and smoking cessation groups. In this supplement, we summarize our understanding of these factors based on interviews with CHWs, primary care providers (PCPs), and study participants. Barriers and facilitators to each of these components of the intervention are outlined in the Online Supplement Table below, which furnishes illustrative quotes associated with each barrier and facilitator as well as implementation recommendations for future iterations of these interventions.

## Provider Education

While a majority of CHWs and PCPs experienced PE to be useful in promoting PCPs’ receptiveness towards prescribing first-line cessation medications to individuals with serious mental illness, some CHWs also described their challenging experiences with PCPs who, despite having received PE, continued to be hesitant about prescribing varenicline due to their perceptions of associated psychiatric risks. Several of the interviewed PCPs did in fact report reluctance to prescribe varenicline due to concerns about psychiatric side effects.

Having an implementation champion in the primary care setting was identified as a facilitator of PCPs’ engagement with PE. For example, PCPs noted being more likely to prioritize smoking cessation and change their TUD prescribing practices when they received this message, or observed a change in practice, from the chief of their service. Several PCPs identified time constraints as the major barrier to PE engagement and implementation. They recommended greater incentives and incorporation of PE into their regular workflow as ways to increase PE attendance and maximize the implementation of best practice guidelines for tobacco cessation.

## Smoking Cessation Groups

CHWs and participants found smoking cessation groups effective in providing goal accountability and positive reinforcement for cessation goals through several strategies: psychoeducation about smoking and health consequences; teaching behavioral change skills; and providing social support and peer accountability, especially through hearing successful quit stories and effective strategies used by group members and guest speakers with lived experience

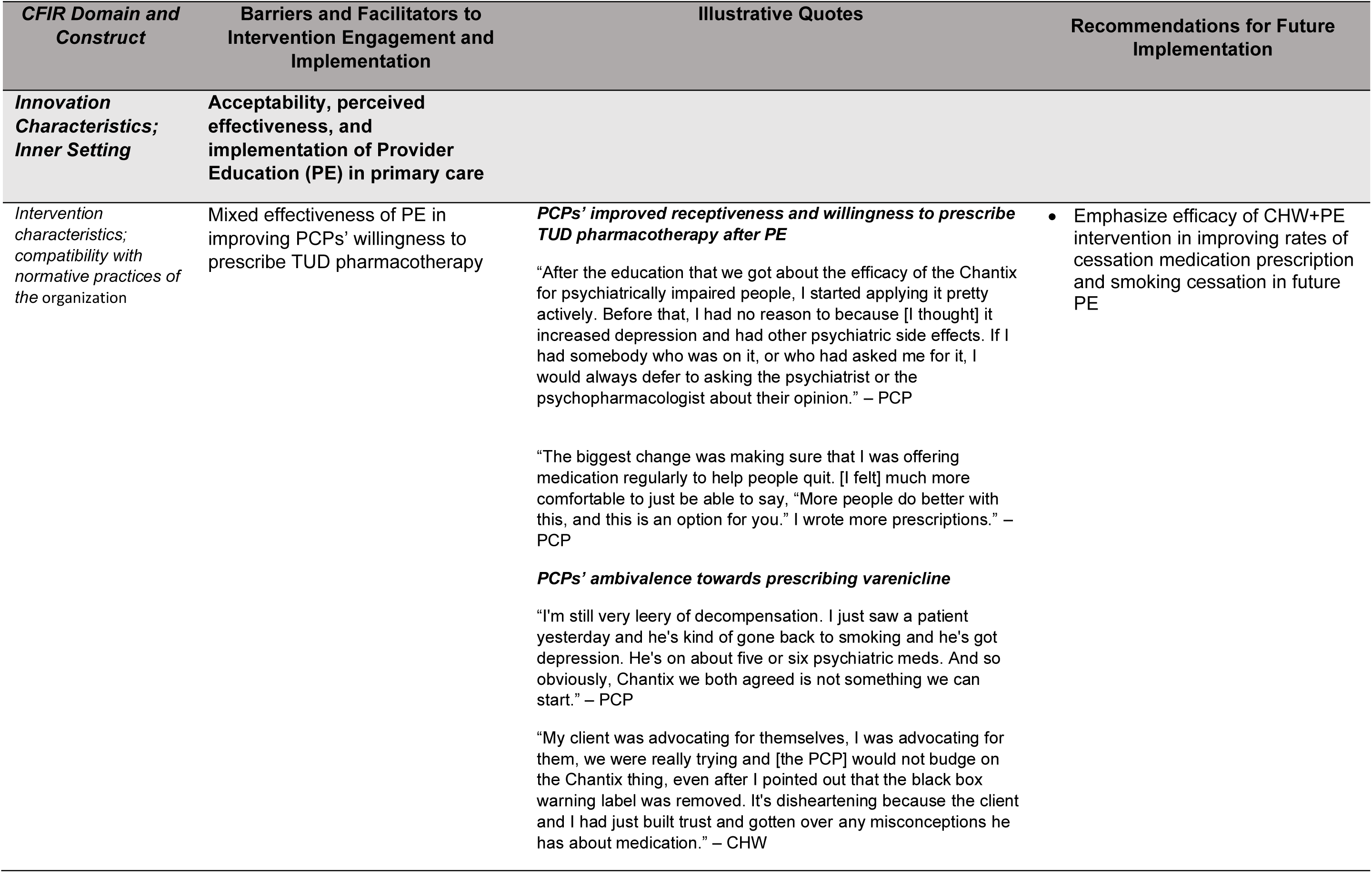
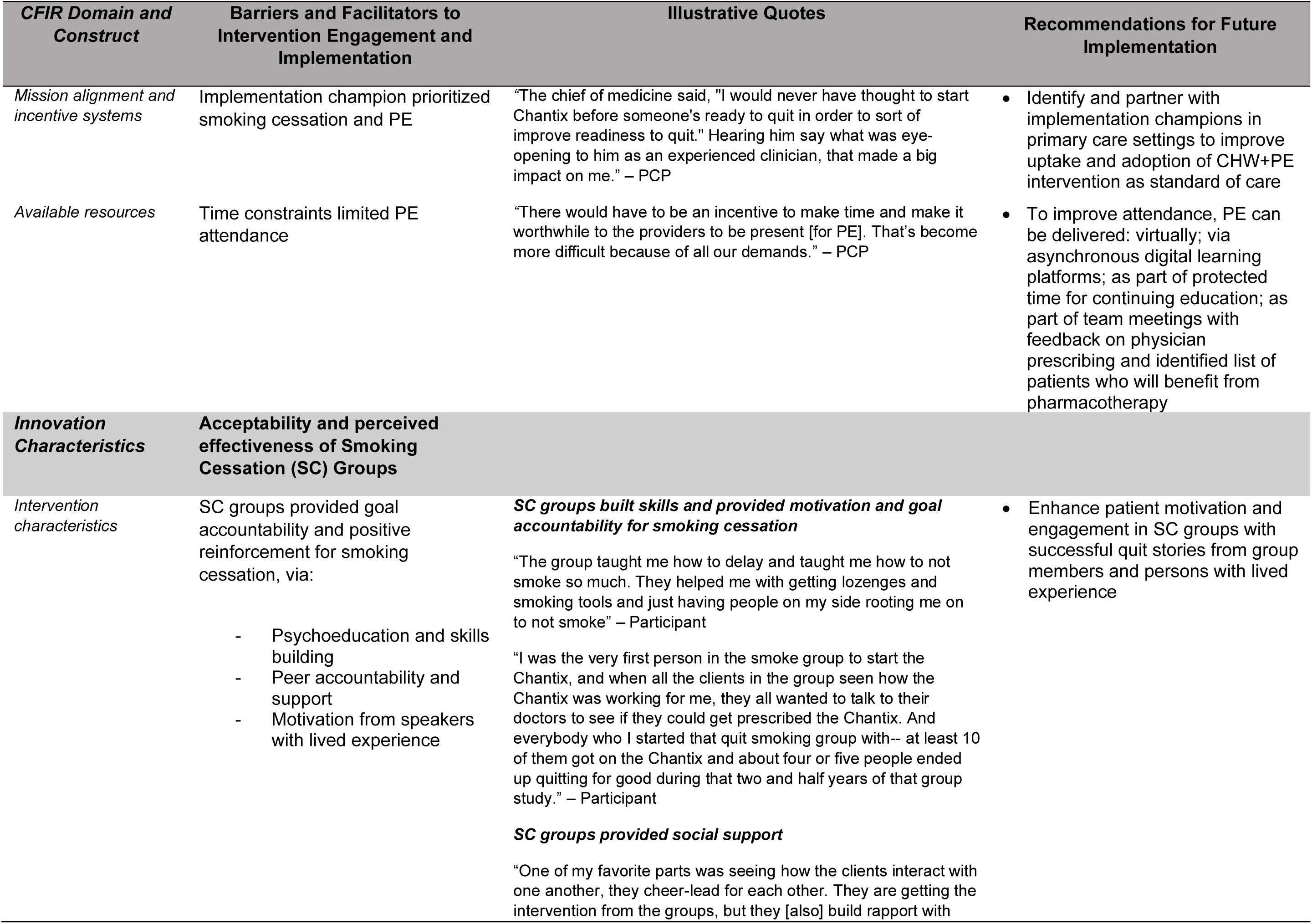
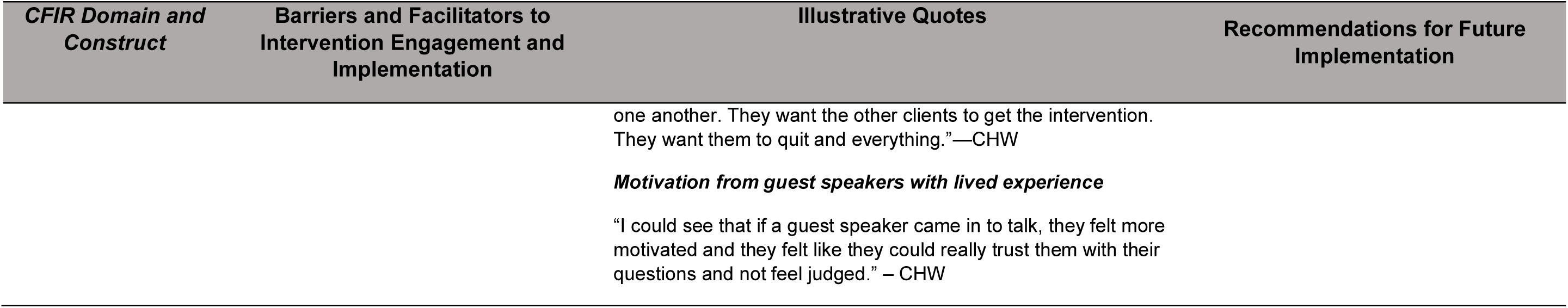
Table of CHW, PCP, and participants’ perceived barriers and facilitators to engagement with and implementation of PE and smoking cessation groups.

| *CFIR Domain and Construct* | *Barriers and Facilitators to Intervention Engagement and Implementation* | *Illustrative Quotes* | *Recommendations for Future Implementation* |
| --- | --- | --- | --- |
| ***Innovation Characteristics; Inner Setting*** | ***Acceptability, perceived effectiveness, and implementation of Provider Education (PE) in primary care*** |  |  |
| *Intervention characteristics; compatibility with normative practices of the organization* | Mixed effectiveness of PE in improving PCPs' willingness to prescribe TUD pharmacotherapy | ***PCPs' improved receptiveness and willingness to prescribe TUD pharmacotherapy after PE***<br>"After the education that we got about the efficacy of the Chantix for psychiatrically impaired people, I started applying it pretty actively. Before that, I had no reason to because [I thought] it increased depression and had other psychiatric side effects. If I had somebody who was on it, or who had asked me for it, I would always defer to asking the psychiatrist or the psychopharmacologist about their opinion." – PCP<br>"The biggest change was making sure that I was offering medication regularly to help people quit. [I felt] much more comfortable to just be able to say, "More people do better with this, and this is an option for you." I wrote more prescriptions." – PCP<br>***PCPs' ambivalence towards prescribing varenicline***<br>"I'm still very leery of decompensation. I just saw a patient yesterday and he's kind of gone back to smoking and he's got depression. He's on about five or six psychiatric meds. And so obviously, Chantix we both agreed is not something we can start." – PCP<br>"My client was advocating for themselves, I was advocating for them, we were really trying and [the PCP] would not budge on the Chantix thing, even after I pointed out that the black box warning label was removed. It's disheartening because the client and I had just built trust and gotten over any misconceptions he has about medication." – CHW | • Emphasize efficacy of CHW+PE intervention in improving rates of cessation medication prescription and smoking cessation in future PE |

ONLINE SUPPLEMENT
| **CFIR Domain and Construct** | **Barriers and Facilitators to Intervention Engagement and Implementation** | **Illustrative Quotes** | **Recommendations for Future Implementation** |
| --- | --- | --- | --- |
| *Mission alignment and incentive systems* | Implementation champion prioritized smoking cessation and PE | "The chief of medicine said, "I would never have thought to start Chantix before someone's ready to quit in order to sort of improve readiness to quit." Hearing him say what was eye-opening to him as an experienced clinician, that made a big impact on me." – PCP | Identify and partner with implementation champions in primary care settings to improve uptake and adoption of CHW+PE intervention as standard of care |
| *Available resources* | Time constraints limited PE attendance | "There would have to be an incentive to make time and make it worthwhile to the providers to be present [for PE]. That's become more difficult because of all our demands." – PCP | To improve attendance, PE can be delivered: virtually; via asynchronous digital learning platforms; as part of protected time for continuing education; as part of team meetings with feedback on physician prescribing and identified list of patients who will benefit from pharmacotherapy |
| **Innovation Characteristics** | **Acceptability and perceived effectiveness of Smoking Cessation (SC) Groups** |  |  |
| *Intervention characteristics* | SC groups provided goal accountability and positive reinforcement for smoking cessation, via:<br>Psychoeducation and skills building<br>Peer accountability and support<br>Motivation from speakers with lived experience | **SC groups built skills and provided motivation and goal accountability for smoking cessation**<br>"The group taught me how to delay and taught me how to not smoke so much. They helped me with getting lozenges and smoking tools and just having people on my side rooting me on to not smoke" – Participant<br>"I was the very first person in the smoke group to start the Chantix, and when all the clients in the group seen how the Chantix was working for me, they all wanted to talk to their doctors to see if they could get prescribed the Chantix. And everybody who I started that quit smoking group with-- at least 10 of them got on the Chantix and about four or five people ended up quitting for good during that two and half years of that group study." – Participant<br>**SC groups provided social support**<br>"One of my favorite parts was seeing how the clients interact with one another, they cheer-lead for each other. They are getting the intervention from the groups, but they [also] build rapport with | Enhance patient motivation and engagement in SC groups with successful quit stories from group members and persons with lived experience |

ONLINE SUPPLEMENT
| *CFIR Domain and Construct* | Barriers and Facilitators to Intervention Engagement and Implementation | Illustrative Quotes | Recommendations for Future Implementation |
| --- | --- | --- | --- |
|  |  | one another. They want the other clients to get the intervention. They want them to quit and everything.”—CHW<br>***Motivation from guest speakers with lived experience***<br>“I could see that if a guest speaker came in to talk, they felt more motivated and they felt like they could really trust them with their questions and not feel judged.” – CHW |  |

